# Acceleration of chemical shift encoding-based water fat MRI for liver proton density fat fraction and T2* mapping using compressed sensing

**DOI:** 10.1101/19000927

**Authors:** Fabian K. Lohöfer, Georgios A. Kaissis, Christina Müller-Leisse, Daniela Franz, Christoph Katemann, Andreas Hock, Johannes M. Peeters, Ernst J. Rummeny, Dimitrios Karampinos, Rickmer F. Braren

**Author notes:** The authors Fabian K. Lohöfer and Georgios A. Kaissis contributed equally to this paper and share the first authorship. Corresponding Author: PD. Dr. med. Rickmer F. Braren, Telephone Nr., E-Mail, Address: Institution Address above.

## Abstract

**Objectives:** To evaluate proton density fat fraction (PDFF) and T2* measurements of the liver with combined parallel imaging (sensitivity encoding, SENSE) and compressed sensing (CS) accelerated chemical shift encoding-based water-fat separation.

**Methods:** Six-echo Dixon imaging was performed in the liver of 89 subjects. The first acquisition variant used acceleration based on SENSE with a total acceleration factor equal to 2.64 (acquisition labeled as SENSE). The second acquisition variant used acceleration based on a combination of CS with SENSE with a total acceleration factor equal to 4 (acquisition labeled as CS+SENSE). Acquisition times were compared between acquisitions and proton density fat fraction (PDFF) and T2*-values were measured and compared separately for each liver segment.

**Results:** Total scan duration was 14.5 sec for the SENSE accelerated image acquisition and 9.3 sec for the CS+SENSE accelerated image acquisition. PDFF and T2* values did not differ significantly between the two acquisitions (*paired Mann-Whitney and paired t-test* P>0.05 in all cases). CS+SENSE accelerated acquisition showed reduced motion artifacts (1.1%) compared to SENSE acquisition (12.3%).

**Conclusion:** CS+SENSE accelerates liver PDFF and T2*mapping while retaining the same quantitative values as an acquisition using only SENSE and reduces motion artifacts.

**Strengths of this study:**

- Compressed sensing allows accelerated imaging with reduction of motion artifacts without alteration of quantitative measurements
- Robust results in fat and iron quantification in a heterogeneous patient cohort

**Limitations of this study:**

- No histopathological validation of the MR findings was performed
- The study was not performed at different field strengths

## Introduction

The non-invasive quantification of fat and iron content in liver tissue is of high clinical significance. For example, Non-Alcoholic Fatty Liver Disease (NAFLD) is the most common cause of chronic liver disease worldwide ^1^, with a prevalence of approximately 30% in adults in the western world ^2^. In patients with NAFLD, liver damage results in hyperferritinemia and hepatic iron accumulation ^3 4^. Both hepatic iron overload and steatosis can result in fibrosis, progress to cirrhosis and therefore carry an increased risk for the development of hepatocellular carcinoma ^5^. Despite availability of non-invasive imaging methods for quantification of hepatic fat and iron content, invasive tissue biopsy and histopathologic visualization of the fat deposition remains the gold standard in detection and quantification of hepatic steatosis and iron overload ^6-12^.

Magnetic resonance imaging (MRI) provides tools for fast, non-invasive quantitative imaging. Specifically, multi-echo gradient-echo acquisitions enable the simultaneous spatially-resolved mapping of the proton density fat fraction (PDFF) and T2*. Liver PDFF has emerged as a method for quantification of intrahepatic fat ^13-17^ with high sensitivity and specificity of 95.0% and 100.0% for the detection of histologic steatosis ^13^. Due to its high diagnostic performance, PDFF is used as a reference modality for other methods of image-based fat quantification like computed tomography ^18^. Likewise, liver T2* mapping has emerged as a method for quantification of intrahepatic iron content ^19^. Chemical shift encoding-based water-fat separation by multi-echo gradient echo acquisition enables the simultaneous accurate and precise quantification of liver PDFF and T2*.

Parallel imaging has been traditionally used to reduce acquisition times, enabling chemical shift encoding-based water-fat separation measurements in a single breath-hold. However, further reduction of the breath-hold duration is highly desirable to avoid motion artefacts and thus improve accuracy of non-invasive fat and iron quantification, especially in patients with difficulty holding their breath. Compressed sensing (CS) allows for acceleration of MRI sequences and has been successfully utilized in various applications ^20-22^. Some methodological works have employed CS for PDFF mapping and applied the technique in small volunteer or patient samples ^23-26^. However, few studies exist on the performance of CS for simultaneous PDFF and T2* mapping in larger patient cohorts.

Therefore, the purpose of this study was the evaluation of the robustness of an CS-accelerated multi-echo gradient echo acquisition for the quantification of hepatic fat and iron content compared to a standard parallel-imaging-accelerated multi-echo gradient echo acquisition in a larger patient cohort.

## Material and Methods

Approval by the institutional ethics committee (180/17S, Ethikkommission der Fakultät für Medizin der Technischen Universität München) was received for the study. The requirement to obtain written informed consent for retrospective data analysis was waived. All analyses were carried out in compliance with the pertinent regulations and requirements.

### Patient and Public Involvement

We did not involve patients or the public in our work

### Patient cohort

We considered 217 datasets of patients who underwent routine clinical liver MRI examination from January 2018 until August 2018 for inclusion in the study. Datasets of patients with primary or secondary/metastatic liver tumors (N=128) were excluded. The final patient cohort consisted of 89 patients (39 males and 50 females). Average patient age was 62.6±16.9 years (range 19-86 years). MRI was performed for the following indications: evaluation of focal pancreatic lesions (n=57), pancreatitis (n=15), evaluation of biliary lesions (n=9) and suspected liver lesions (n=8).

### Data acquisition

Two variants of multi-echo gradient-echo imaging for performing chemical shift encoding-based water-fat separation were performed sequentially on each patient at a 3 T MRI scanner (*Philips Ingenia Elition X; Philips Medical Systems*, Best, The Netherlands). The two acquisitions were based on a spoiled gradient echo sequence using bipolar gradient readouts. The first acquisition variant used acceleration based on SENSE with a total acceleration factor equal to 2.64 (acquisition labeled as SENSE). The second acquisition variant used acceleration based on a combination of CS with SENSE with a total acceleration factor equal to 4 (acquisition labeled as CS+SENSE). The relevant scan parameters are listed in *Table 1*.

**Table 1:** Scan parameters

|  | **Acquisition**<br>**with SENSE** | **Acquisition with**<br>**CS+SENSE** |
| --- | --- | --- |
| **Gating** | Breath-hold | Breath-hold |
| **Acquisition duration (s)** | 14.5 | 9.3 |
| **Acceleration factor** | $2.2 \times 1.2 = 2.64$ | 4 |
| **FOV (mm) FH; RL; AP** | 150; 400; 300 | 150; 400; 300 |
| **Acquisition voxel size (mm) FH; RL; AP** | 6; 3; 2 | 6; 3; 2 |
| **Reconstruction voxel size (mm)**<br>**FH/RL/AP** | 6; 1.14; 1.14 | 6; 1.14; 1.14 |
| **Fast imaging mode** | none | none |
| **TE<sub>eff</sub>/TE<sub>equiv</sub> (ms)** |  |  |
| **Act. TR (ms)** | 7.8 | 7.8 |
| **Act. TE (ms)** | 1.35 | 1.35 |
| **Flip angle (°)** | 3 | 3 |

The CS+SENSE technique used in the present work was based on the combination of SENSE and CS, labelled as Compressed SENSE or C-SENSE. The technique uses the coil sensitivity information from a SENSE calibration scan and randomly undersamples both the central and outer part of *k*-space, following a smooth sampling density as moving from the center to outer parts of k-space. The acquisition and reconstruction were based on the vendor’s implementation (Compressed SENSE, Philips Healthcare). A single CS acceleration factor was defined for the CS+SENSE acquisition variant and the sampled k-space pattern (central and outer part) was defined based on the vendor’s implementation. In order to maintain a balance between noise reduction and data consistency for CS, an iterative L1-minimization reconstruction technique, forcing data fidelity, and image sparsity in the wavelet domain, was used.

Complex multi-echo gradient-echo images were generated after the SENSE and CS+SENSE reconstructions and provided as input to the fat quantification routine provided by the vendor (*mDixon Quant*, Philips Healthcare). Specifically, after phase correction, a complex-based water-fat decomposition was performed using a single T_2_* correction and a pre-calibrated fat spectrum accounting for the presence of the multiple peaks in the fat spectrum. A seven-peak fat spectrum model was employed ^27^. The PDFF map was computed as the ratio of the fat signal over the sum of fat and water signals.

### Image analysis

Images were reviewed by two abdominal radiologists under standardized radiological reporting room conditions. Circular regions of interest (ROI) with a diameter of 15mm were manually drawn in each liver segment in consensus, avoiding large portal vein and hepatic vein branches (*Figure 1)*. T2* and PDFF maps were reviewed and mean ROI values and standard deviations were extracted. The software used was *Sectra IDS7 (*Linköping, Sweden). **Motion artifacts** were rated using a 4-point *Likert* scale as *1=image not diagnostic because of artifacts; 2=major artifacts; 3=minor artifacts; 4=no artifacts*.

**Figure 1:**
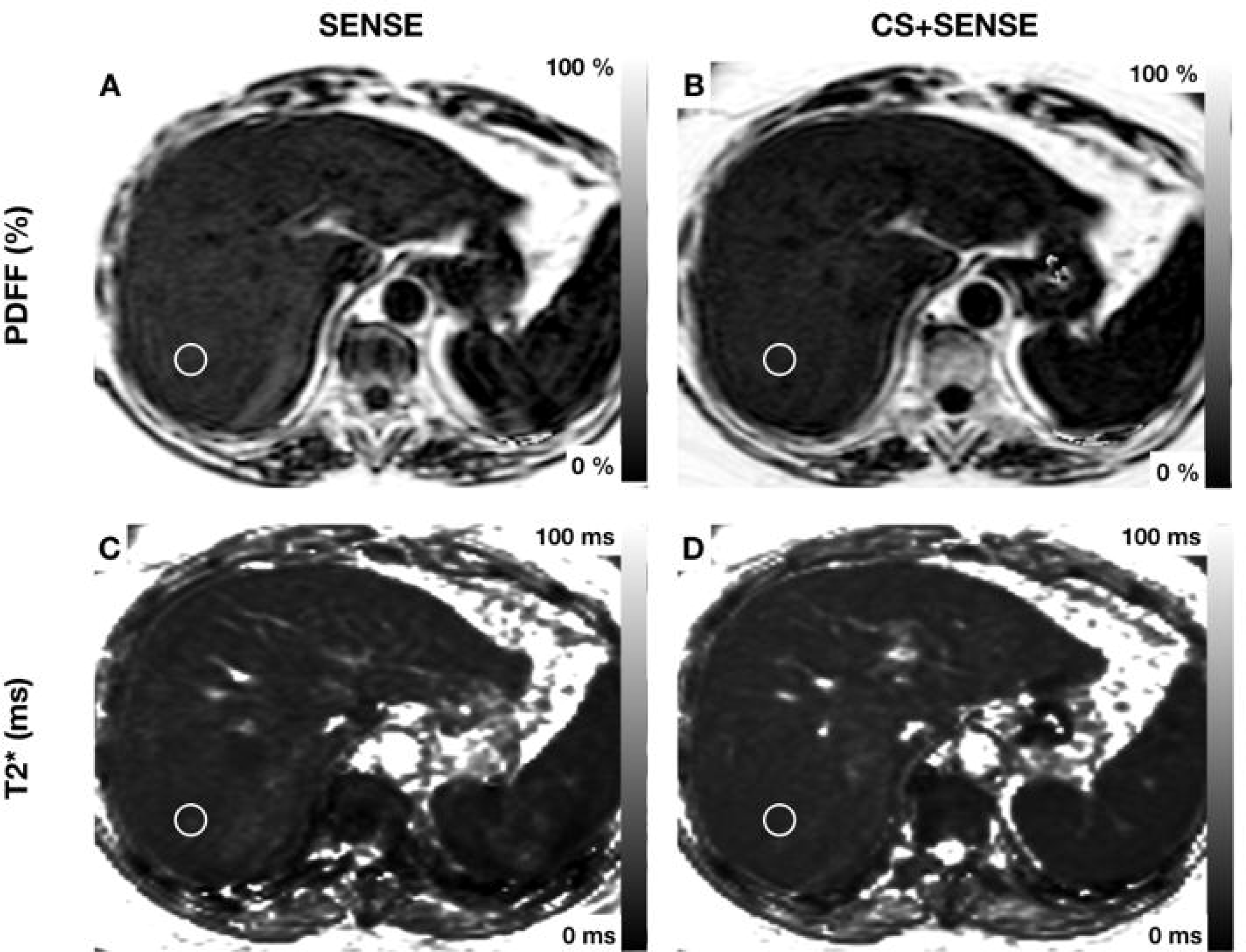
MRI was performed for evaluation of cystic pancreatic lesion. Exemplary region of interest is drawn in segment VII (1.5cm). PDFF: acquisition with SENSE 11.1±2.4% (A); acquisition with CS+SENSE 10.8±1.5% (B); T2*: acquisition with SENSE 9.35±0.6ms (C); acquisition with CS+SENSE 9.28±0.5ms (D)

### Statistical analysis

Variables were tested for normal distribution using the *D’Agostino-Pearson omnibus K2 test. Student’s t-test* was used for mean comparisons of normally distributed variables. The *Wilcoxon* test was used for mean comparisons of variables without normal distribution. All analyses were performed using *Prism* Version 7 (*GraphPad Software*). A two-tailed P-value below 0.05 was considered statistically significant.

## Results

The use of CS+SENSE accelerated image acquisition by 35% compared to the SENSE acquisition (from 14.5 to 9.3 seconds). Mean PDFF values ranged from 4.94 ± 5.8% (liver segment IVb) to 6.37 ± 6.65% (liver segment VII) in the acquisition with SENSE and from 5.00 ± 6.32% (liver segment IVb) to 6.28 ± 6.10% (liver segment VII) in the acquisition with CS+SENSE. Results did not differ significantly between the two acquisitions. All values are shown in *Table 2*. Mean PDFF was significantly higher in the right liver lobe compared to the left in both acquisitions (right lobe: 6.04 ± 6.36%; left lobe: 5.24 ± 5.69%; P=0.03; acquisition with SENSE and right lobe: 5.96 ± 6.30%; left lobe: 5.16 ± 5.74%; P=0.02; acquisition with CS+SENSE).

**Table 2:** PDFF mean values in % with standard deviation, no significant differences were seen between the two acquisitions. Wilcoxon test (no normal distribution); Patients: n=89

| Liver Segment | PDFF (%) in acquisition with SENSE | PDFF (%) in acquisition with CS+SENSE | p |
| --- | --- | --- | --- |
| **I** | 5.36±4.97 | 5.11±4.61 | 0.1624 |
| **II** | 5.53±6.23 | 5.31±5.90 | 0.6088 |
| **III** | 5.29±5.83 | 5.28±6.03 | 0.9100 |
| **IVa** | 5.08±5.68 | 5.12±5.79 | 0.5262 |
| **IVb** | 4.94±5.80 | 5.00±6.32 | 0.8236 |
| **V** | 5.53±6.73 | 5.50±6.73 | 0.6557 |
| **VI** | 6.19±5.93 | 6.05±5.76 | 0.1847 |
| **VII** | 6.37±6.19 | 6.28±6.10 | 0.7536 |
| **VIII** | 6.05±6.65 | 6.00±6.66 | 0.9425 |

Mean T2* values ranged from 20.54±7.05 ms (liver segment II) to 23.42±7.43 ms (liver segment V) in the acquisition with SENSE and from 21.29±8.24 ms (liver segment II) to 23.30±8.60 ms (liver segment IVb) in the acquisition with CS+SENSE. T2* values did not differ significantly between the two acquisitions. T2* values with inter-patient standard deviation and results of the mean comparison are shown in *Table 3*. T2* values showed no significant difference between the right and left liver lobe in both acquisitions (right lobe: 22.60 ± 7.28ms; left lobe: 21.70 ± 7.14ms; P=0.08 in the acquisition with SENSE; right lobe: 22.65 ± 7.31ms; left lobe: 22.14 ± 7.94ms; P=0.35; in the acquisition with CS+SENSE). Images acquired with SENSE showed minor motion artifacts in 10.1% (n=9) and major motion artifacts in 2.2% (n=2) of the cases. Images acquired with CS+SENSE showed minor motion artifacts in 1.1% (n=1) of all cases. One exemplary case with hepatic steatosis, sparing segment I is shown in *Figure 2. Figure 3* shows a patient with segmental steatosis. A T2 weighted image and a CT-image, acquired in the portal venous phase are shown as a comparison. *Figure 4* shows a case of a patient with hepatic iron overload due to hemosiderosis.

**Table 3:** T2* mean values in ms with standard deviation no significant differences were seen between the two acquisitions. Paired t test (normal distribution); Patients: n=89

| Liver Segment | T2* (ms) in acquisition with SENSE | T2* (ms) in acquisition with CS+SENSE | p |
| --- | --- | --- | --- |
| **I** | 22.25±7.70 | 23.04±7.67 | 0.1578 |
| **II** | 20.54±7.05 | 21.29±8.24 | 0.2096 |
| **III** | 20.96±6.98 | 21.38±7.85 | 0.3541 |
| **IVa** | 21.59±6.48 | 21.68±7.22 | 0.8428 |
| **IVb** | 23.15±7.30 | 23.30±8.60 | 0.7841 |
| **V** | 23.42±7.43 | 23.15±7.46 | 0.3549 |
| **VI** | 22.91±6.94 | 22.97±7.21 | 0.8399 |
| **VII** | 22.05±7.38 | 22.18±7.34 | 0.7391 |
| **VIII** | 22.05±6.76 | 22.32±7.31 | 0.4264 |

**Figure 2:**
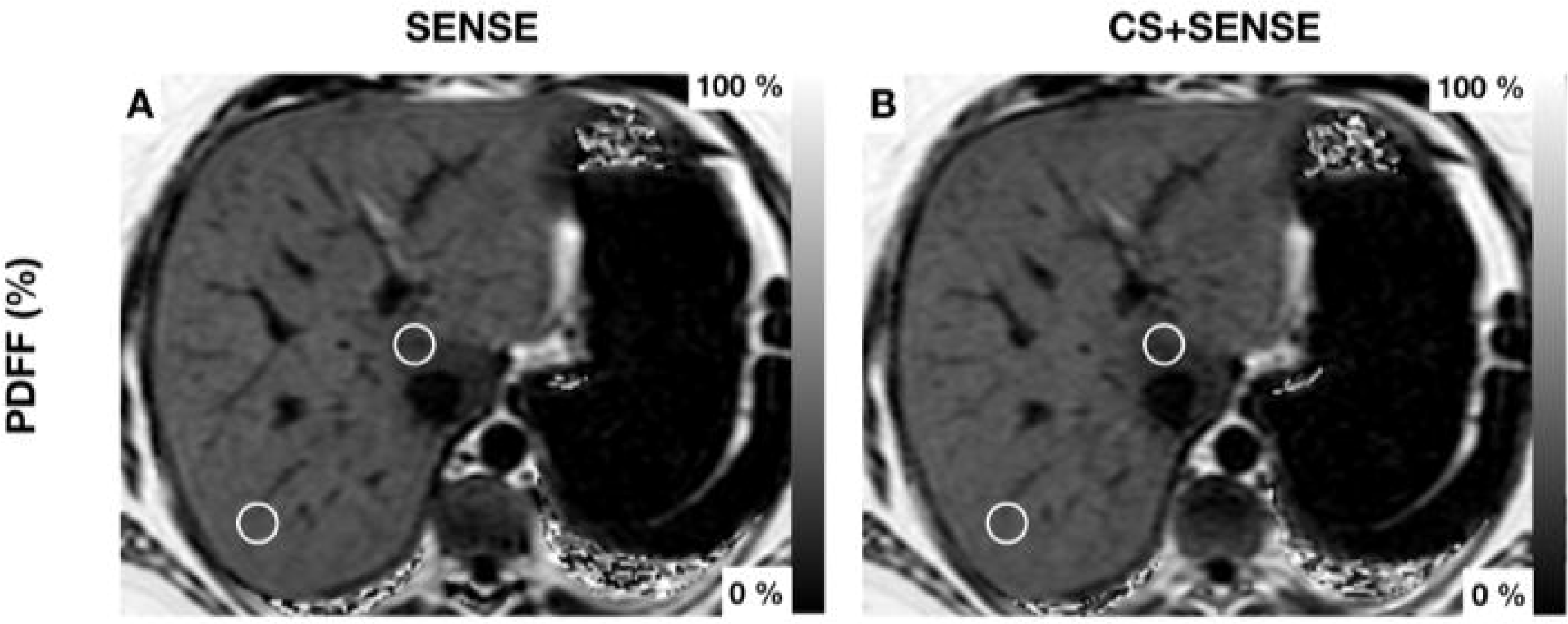
Patient with hepatic steatosis with focal fatty sparing of Segment I. Exemplary region of interest are drawn in Segments I and VII (1.5cm). PDFF Segment I: acquisition with SENSE 25.9±3.8% (A); acquisition with CS+SENSE CS4 25.8±4.5% (B) PDFF Segment VII: acquisition with SENSE 32.2±2.0% (A); acquisition with CS+SENSE 33.2±1.6% (B)

**Figure 3:**
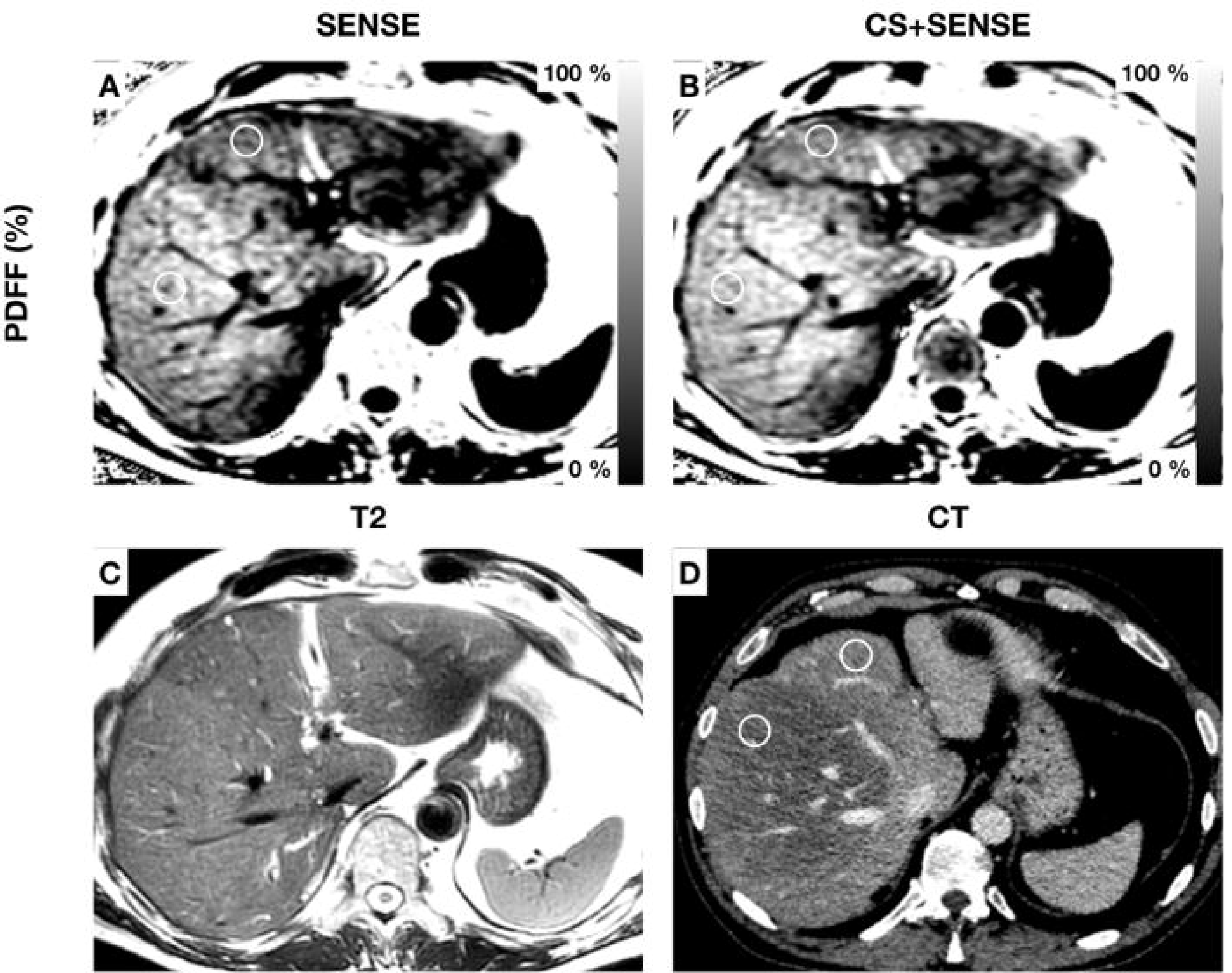
Patient with segmental hepatic steatosis. Exemplary regions of interest are drawn in segments IVa and V (diameter 1.5cm). PDFF Segment IVa: acquisition with SENSE 25.7±1.5% (A); acquisition with CS+SENSE 26.9±2.0% (B) PDFF Segment V: acquisition with SENSE 31.6±1.8% (A); acquisition with CS+ SENSE 32.6±2.2 (B) A T2-weighted image (C) and a CT image in the venous contrast phase (D) are shown for comparison. In CT Hounsfield units are 87±13 in Segment IVa and 42±16 in Segment V

**Figure 4:**
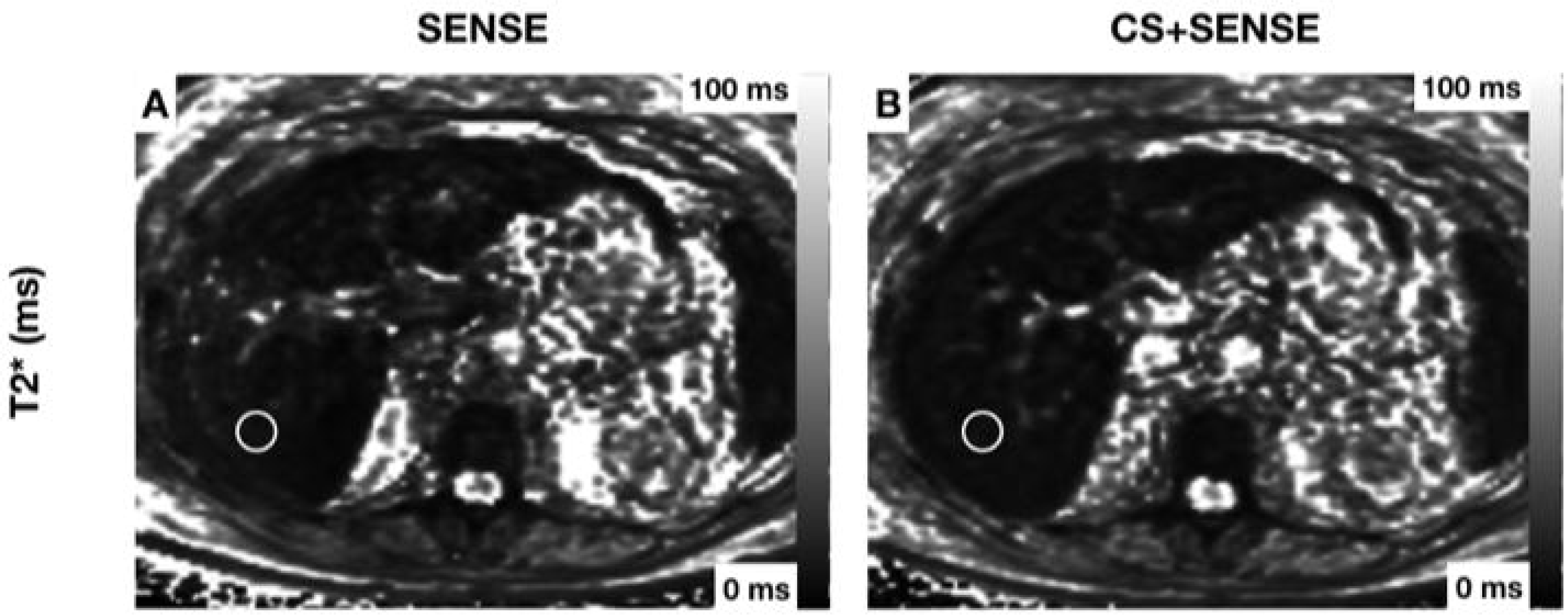
Patient with hemosiderosis and severe reduction of T2* Exemplary region of interest is drawn in segment VII (1.5cm). T2*: acquisition with SENSE 4.03±0.8ms (A); acquisition with CS+SENSE 5.56±1.0ms (B)

## Discussion

The data presented in this study show comparability of quantitative PDFF and T2* measurements acquired with compressed sensing (CS)-accelerated chemical shift encoding-based water-fat separation. Our results demonstrate a significantly higher PDFF in the right lobe of the liver, which is in accordance with other studies ^28 29^, and no spatial dependence of liver T2* values.

The most important finding of the presented data is the agreement of the quantitative PDFF and T2* imaging results between the two acquisitions. Several studies have shown high accuracy of MRI-based imaging techniques for the non-invasive quantification of fat and iron content of the liver ^13 14 29^. Due to their widespread availability, chemical shift encoding-based water-fat separation techniques can be used as a fast screening for NAFLD or disorders of iron metabolism. Given the increase of medical imaging in the last years ^30^, the acquisition time is an essential factor. In our study, CS+SENSE was able to accelerate image acquisition of liver PDFF and T2* mapping by 35% to a scan time of only 9.3 seconds. In a liver segment-based comparison between the acquisition with CS+SENSE and the acquisition with SENSE, no significant differences were detected in the quantitative parameters PDFF and T2*. Thus, reduction of scan time lead to a reduction of motion artifacts and did not lead to changes in quantitative parameters, rendering the presented chemical shift encoding-based water-fat separation technique combined with CS+SENSE a sequence with an excellent applicability in routine MRI liver examinations. The widespread availability could allow an application for large studies on hepatic steatosis.

Previous methodological works applied CS for the joint problem of image reconstruction and water-fat separation ^23-25^. The present work estimates the PDFF and T2* maps in two steps: it first applies the CS+SENSE reconstruction for the reconstruction of the multi-echo complex images and then applies water-fat separation on the reconstructed water-fat images, as also previously performed ^26 31^. Although higher acceleration factors can be achieved by solving the joint step instead of solving the problem in two steps, the present study shows that a prospective CS+SENSE accelerated acquisition using the two-step approach already results in reliable PDFF and T2* maps in the presently studied patient cohort.

Our study has some limitations. First, the study was only performed in one center and on one scanner to show the feasibility of the accelerated acquisition with CS+SENSE. We did not test the two acquisitions at different field strengths. Second, histologic correlation was not performed. However, this was not within the scope of the current study that aimed at showing robustness of quantitative parameters when using CS acceleration. In addition, several previous studies have already shown high sensitivity and specificity in liver quantitative imaging with chemical shift encoding based water-fat separation techniques accounting for the same confounding factors 13 32.

In conclusion, the acceleration of chemical shift encoding-based water-fat separation using compressed sensing results in comparable quantitative measurements of hepatic fat and iron content with reduced breath-hold intervals, leading to reduced motion artifacts and making chemical shift encoding-based water-fat separation a fast and precise non-invasive tool in quantitative liver imaging.

## Data Availability

The data generated during the study is available from the corresponding author on reasonable request by a qualified individual or third-party, subject to authorization of pertinent regulatory bodies.

## Author contribution statement (CREDIT Taxonomy)

Conceptualization: FL, GK, RB

Data Curation: FL, GK, CML

Formal Analysis: FL, GK

Funding Acquisition: N.A.

Investigation: FL, GK, CM

Methodology: FL, GK, RB

Project Administration: FL, RB, DK

Resources: DK, AH, CK, JP

Software: GK, DK

Supervision: RB, DK, ER

Validation: FL, GK

Visualization: FL, GK

Writing- Original Draft: FL, GK

Writing- Review and Editing: FL, GK, DF, RB

The authors Fabian K. Lohöfer (FL) and Georgios A. Kaissis (GK) contributed equally to this paper and share the first authorship

## References

1. Fraser A, Harris R, Sattar N, et al. Alanine aminotransferase, gamma-glutamyltransferase, and incident diabetes: the British Women’s Heart and Health Study and meta-analysis. Diabetes Care 2009;32(4):741–50. doi: 10.2337/dc08-1870

2. Zelber-Sagi S, Nitzan-Kaluski D, Halpern Z, et al. Prevalence of primary non-alcoholic fatty liver disease in a population-based study and its association with biochemical and anthropometric measures. Liver Int 2006;26(7):856–63. doi: 10.1111/j.1478-3231.2006.01311.x

3. Valenti L, Fracanzani AL, Dongiovanni P, et al. Iron depletion by phlebotomy improves insulin resistance in patients with nonalcoholic fatty liver disease and hyperferritinemia: evidence from a case-control study. Am J Gastroenterol 2007;102(6):1251–8. doi: 10.1111/j.1572-0241.2007.01192.x [Published Online First: 2007/03/30]

4. Valenti L, Fracanzani AL, Bugianesi E, et al. HFE genotype, parenchymal iron accumulation, and liver fibrosis in patients with nonalcoholic fatty liver disease. Gastroenterology 2010;138(3):905–12. doi: 10.1053/j.gastro.2009.11.013 [Published Online First: 2009/11/26]

5. Olivieri NF, Brittenham GM. Iron-chelating therapy and the treatment of thalassemia. Blood 1997;89(3):739–61.

6. Adams LA, Angulo P, Lindor KD. Nonalcoholic fatty liver disease. CMAJ 2005;172(7):899–905. doi: 10.1503/cmaj.045232

7. Kleiner DE, Brunt EM, Van Natta M, et al. Design and validation of a histological scoring system for nonalcoholic fatty liver disease. Hepatology 2005;41(6):1313–21. doi: 10.1002/hep.20701

8. Sanyal AJ, American Gastroenterological A. AGA technical review on nonalcoholic fatty liver disease. Gastroenterology 2002;123(5):1705–25.

9. Brunt EM, Janney CG, Di Bisceglie AM, et al. Nonalcoholic steatohepatitis: a proposal for grading and staging the histological lesions. Am J Gastroenterol 1999;94(9):2467–74. doi: 10.1111/j.1572-0241.1999.01377.x

10. Sarigianni M, Liakos A, Vlachaki E, et al. Accuracy of magnetic resonance imaging in diagnosis of liver iron overload: a systematic review and meta-analysis. Clin Gastroenterol Hepatol 2015;13(1):55–63 e5. doi: 10.1016/j.cgh.2014.05.027 [Published Online First: 2014/07/06]

11. Nash S, Marconi S, Sikorska K, et al. Role of liver biopsy in the diagnosis of hepatic iron overload in the era of genetic testing. Am J Clin Pathol 2002;118(1):73–81. doi: 10.1309/4A4U-N4GL-DRP3-EQPD

12. Urru SA, Tandurella I, Capasso M, et al. Reproducibility of liver iron concentration measured on a biopsy sample: a validation study in vivo. Am J Hematol 2015;90(2):87–90. doi: 10.1002/ajh.23878

13. Yokoo T, Bydder M, Hamilton G, et al. Nonalcoholic fatty liver disease: diagnostic and fat-grading accuracy of low-flip-angle multiecho gradient-recalled-echo MR imaging at 1.5 T. Radiology 2009;251(1):67–76. doi: 10.1148/radiol.2511080666

14. Reeder SB, Sirlin CB. Quantification of liver fat with magnetic resonance imaging. Magn Reson Imaging Clin N Am 2010;18(3):337-57, ix. doi: 10.1016/j.mric.2010.08.013

15. Reeder SB, Cruite I, Hamilton G, et al. Quantitative assessment of liver fat with magnetic resonance imaging and spectroscopy. J Magn Reson Imaging 2011;34(4):729–49. doi: 10.1002/jmri.22580

16. Reeder SB, Hu HH, Sirlin CB. Proton density fat-fraction: a standardized MR-based biomarker of tissue fat concentration. J Magn Reson Imaging 2012;36(5):1011–4. doi: 10.1002/jmri.23741

17. Hu HH, Kan HE. Quantitative proton MR techniques for measuring fat. NMR Biomed 2013;26(12):1609–29. doi: 10.1002/nbm.3025

18. Zhang Y, Wang C, Duanmu Y, et al. Comparison of CT and magnetic resonance mDIXON-Quant sequence in the diagnosis of mild hepatic steatosis. Br J Radiol 2018;91(1091):20170587. doi: 10.1259/bjr.20170587

19. Hernando D, Levin YS, Sirlin CB, et al. Quantification of liver iron with MRI: state of the art and remaining challenges. J Magn Reson Imaging 2014;40(5):1003–21. doi: 10.1002/jmri.24584

20. Feng L, Benkert T, Block KT, et al. Compressed sensing for body MRI. J Magn Reson Imaging 2017;45(4):966–87. doi: 10.1002/jmri.25547

21. Fushimi Y, Fujimoto K, Okada T, et al. Compressed Sensing 3-Dimensional Time-of-Flight Magnetic Resonance Angiography for Cerebral Aneurysms: Optimization and Evaluation. Invest Radiol 2016;51(4):228–35. doi: 10.1097/RLI.0000000000000226

22. Zhu L, Wu X, Sun Z, et al. Compressed-Sensing Accelerated 3-Dimensional Magnetic Resonance Cholangiopancreatography: Application in Suspected Pancreatic Diseases. Invest Radiol 2018;53(3):150–57. doi: 10.1097/RLI.0000000000000421

23. Doneva M, Bornert P, Eggers H, et al. Compressed sensing for chemical shift-based water-fat separation. Magn Reson Med 2010;64(6):1749–59. doi: 10.1002/mrm.22563

24. Sharma SD, Hu HH, Nayak KS. Accelerated T2*-compensated fat fraction quantification using a joint parallel imaging and compressed sensing framework. J Magn Reson Imaging 2013;38(5):1267–75. doi: 10.1002/jmri.24034

25. Wiens CN, McCurdy CM, Willig-Onwuachi JD, et al. R2*-corrected water-fat imaging using compressed sensing and parallel imaging. Magn Reson Med 2014;71(2):608–16. doi: 10.1002/mrm.24699

26. Mann LW, Higgins DM, Peters CN, et al. Accelerating MR Imaging Liver Steatosis Measurement Using Combined Compressed Sensing and Parallel Imaging: A Quantitative Evaluation. Radiology 2016;278(1):247–56. doi: 10.1148/radiol.2015150320

27. Ren J, Dimitrov I, Sherry AD, et al. Composition of adipose tissue and marrow fat in humans by 1H NMR at 7 Tesla. J Lipid Res 2008;49(9):2055–62. doi: 10.1194/jlr.D800010-JLR200

28. Hua B, Hakkarainen A, Zhou Y, et al. Fat accumulates preferentially in the right rather than the left liver lobe in non-diabetic subjects. Dig Liver Dis 2018;50(2):168–74. doi: 10.1016/j.dld.2017.08.030

29. Fazeli Dehkordy S, Fowler KJ, Mamidipalli A, et al. Hepatic steatosis and reduction in steatosis following bariatric weight loss surgery differs between segments and lobes. European radiology 2018 doi: 10.1007/s00330-018-5894-0

30. Smith-Bindman R, Miglioretti DL, Larson EB. Rising use of diagnostic medical imaging in a large integrated health system. Health Aff (Millwood) 2008;27(6):1491–502. doi: 10.1377/hlthaff.27.6.1491

31. Hollingsworth KG, Higgins DM, McCallum M, et al. Investigating the quantitative fidelity of prospectively undersampled chemical shift imaging in muscular dystrophy with compressed sensing and parallel imaging reconstruction. Magn Reson Med 2014;72(6):1610–9. doi: 10.1002/mrm.25072

32. Serai SD, Smith EA, Trout AT, et al. Agreement between manual relaxometry and semi-automated scanner-based multi-echo Dixon technique for measuring liver T2* in a pediatric and young adult population. Pediatr Radiol 2018;48(1):94–100. doi: 10.1007/s00247-017-3990-y

